# Characterizing Documented Psychosocial Stressors in Pediatric Psychiatric Emergencies with an Open-Weight Large Language Model

**DOI:** 10.64898/2026.06.08.26354931

**Authors:** Carson S Hartlage, Erika Rasnick Manning, Jonathan Bernard, Sia Vaish, James Gray, Melissa Young, Teresa Pestian, Patricia Tachinardi, Eneida A Mendonca, Alonzo T Folger, Cole Brokamp

## Abstract

**Objective:** To evaluate whether a locally hosted open-weight large language model (LLM) can extract documented psychosocial factors from pediatric psychiatric intake notes and apply validated extraction to a large emergency psychiatry cohort.

**Materials and Methods:** We identified emergency department presentations at Cincinnati Children’s Hospital Medical Center from January 1, 2016, through December 31, 2024, among patients <18 years with psychiatric billing diagnoses. Using full-text intake notes, gpt-oss:120b classified peer conflict, sleep disruption, and school-related academic, attendance, and disciplinary issues as detected, negated, or indeterminate. Four human raters independently reviewed 50 notes. We compared Fleiss’ κ among humans alone versus humans plus the LLM, assessed repeated-query stability across 50 independent calls per note, and applied the workflow to all eligible notes.

**Results:** Among 37,315 eligible admissions, 22,284 had eligible intake notes; 22,270 produced parseable JSON. In detected-vs-not-detected coding, human-plus-LLM reliability did not differ significantly from human-only reliability across measures (human κ, 0.71–0.94; human-plus-LLM κ, 0.70–0.93). Stability was associated with human agreement: mean LLM-human agreement increased from 42.6% for classifications with <80% stability to 82.7% for 100% stability (Pearson r=0.36). Full-cohort extraction showed frequent and overlapping documented factors: sleep disruption was most frequently detected (57.7%), followed by peer conflict (47.2%), academic issues (43.4%), disciplinary issues (43.3%), and attendance issues (16.9%).

**Discussion:** Agreement varied by construct and was strongest when repeated model outputs were stable.

**Conclusion:** Locally hosted open-weight LLMs can support scalable structured extraction of documented psychosocial factors from pediatric psychiatric intake notes after local validation.

## BACKGROUND AND SIGNIFICANCE

Pediatric psychiatric emergency presentations have increased substantially.[1] In national US data, emergency department visits for children with mental health disorders increased by 60% from 2007 to 2016, including a marked increase in visits for deliberate self-harm.[2] Mental health emergency visits and revisits have also increased in children’s hospitals.[3] The crisis in pediatric mental health has created an urgency for scalable methods to characterize the clinical and psychosocial context of these encounters and to inform risk stratification and follow-up planning.

Pediatric psychiatric assessments are conducted by clinicians during an emergency encounter and contain clinically important information about the circumstances surrounding a mental health crisis. Assessments are stored as free text data and often document family processes and proximal psychosocial stressors, including interpersonal, school, and family events, that may shape risk, intervention planning, disposition, and downstream outcomes.[4] This contextual information is central to youth crisis assessment; pediatric emergency mental health screening tools explicitly incorporate domains such as home, education, activities and peers, suicidality, emotions and behavior, and discharge resources.[5]

However, much of this information is stored as unstructured narrative text or in semi-structured flowsheets rather than in stable, analysis-ready fields. Structured electronic health records (EHR) fields are useful when available, but they do not capture the full psychosocial context of pediatric psychiatric emergencies. Reuse of EHR data for research requires task-specific assessment of data quality dimensions such as completeness, correctness, concordance, plausibility, and currency.[6] For social and behavioral determinants, structured EHR formats remain inconsistently implemented, and relevant information is often absent from structured fields or documented only in narrative text.[7] Data captured in clinical documentation content can also be site-specific, may lack a well-defined data dictionary, and can change over time or across service lines as clinical workflows and EHR templates evolve. As a result, observations of the same phenomenon may be represented inconsistently, while details such as school issues, bullying, and other recent interpersonal stressors may appear only in free text. This limits the ability of observational EHR-based research to capture the full contextual factors contributing to pediatric psychiatric emergencies.

Natural language processing (NLP) offers a path to transform these narrative assessments into structured variables at scale. Proprietary clinical NLP systems, including named-entity recognition tools, can support extraction from clinical notes but may be costly, less flexible for locally defined concepts, or difficult to adapt to institution-specific documentation. Increasingly accessible open-weight large language models (LLM) provide a quality alternative approach because the tools can be hosted within secure institutional environments, queried with locally developed prompts, and asked to return structured data outputs. LLMs are increasingly trained and evaluated for reason-oriented language tasks, which can use contextual reasoning to interpret variable, narrative documentation rather than relying primarily on fixed keywords or rule-based patterns. Furthermore, mixture-of-experts architectures have enabled scalable computation by utilizing specialized subnetworks, called experts, that are activated only when needed. This effectively allows for a very large total parameter count while using much less compute per token than a dense model of the same size.

Recent clinical studies have evaluated open-source or local large language models for clinical information extraction, including structured output generation, while also demonstrating the need for task-specific validation because performance varies across extraction targets and models may struggle with output format consistency or hallucination.[8,9] Psychiatry- and pediatric-relevant examples are emerging: large language models have been compared with rule-based NLP for extracting social support and social isolation from psychiatry notes,[10] used to estimate dimensional psychopathology from adult discharge notes [11] and pediatric psychiatric clinical notes,[12] and reviewed across pediatric clinical text applications.[13]

Despite this progress, important gaps remain for EHR-based pediatric psychiatric research. Much existing clinical LLM extraction work has focused on discrete entities, short text spans, or sentence-level classification, and has often relied on concepts that are directly documented in the note or on task-specific model fine-tuning. Less is known about whether a locally hosted open-weight model can, without fine-tuning, interpret complete psychiatric intake assessments and return accessible structured outputs for fine-grained psychosocial constructs that are clinically relevant but variably documented, such as peer conflict, sleep disruption, and specific school-related difficulties. Demonstrating this capability is important because zero-shot, note-level extraction can reduce the need for large, annotated training sets, preserve contextual information distributed across long notes, and make locally defined measures easier for clinical and non-informatics collaborators to specify, inspect, and reuse through structured schemas. This study addresses this gap by validating an on-premises open-weight LLM against human reviewers and then applies the same workflow to more than 25,000 psychiatric intake notes, enabling large-scale characterization of documented psychosocial stressors and their intersections during pediatric psychiatric emergency encounters.

## OBJECTIVE

This study evaluates whether an on-premises open-weight large language model can reliably extract five psychosocial variables from pediatric psychiatric intake notes from a large pediatric academic medical center. We compare model outputs and stability across repeated queries with human ratings, then extract from all notes to classify the prevalence of select social and behavioral determinants across all noted emergency psychiatric intakes.

## MATERIALS AND METHODS

We identified admissions through the emergency department by patients aged less than 18 years at Cincinnati Children’s Hospital Medical Center (CCHMC) from January 1, 2016, through December 31, 2024. CCHMC is a 629-bed pediatric academic medical center and is the dominant source of pediatric hospital care in Hamilton County, Ohio. CCHMC is the largest inpatient pediatric mental health provider in the country and operates the College Hill Campus, a pediatric behavioral health campus that provides inpatient, outpatient, and residential behavioral health services. Admissions were considered if they had at least one billing diagnosis ICD-10 categorized in a psychiatric exacerbation category defined by the Pediatric Clinical Classification System (PECCS). Because PECCS categories are intentionally broad, we worked with a child psychiatrist to apply an a priori code-level refinement to retain only diagnoses with face validity for acute pediatric emergency mental health encounters. We excluded selected F-codes that represented mental disorders due to known physiological or medical conditions (e.g., F06.4, F06.30, F06.0), chronic or outpatient-oriented conditions unlikely to drive an ED visit (e.g., F30.1, F21), residual or nonspecific diagnostic categories (e.g. F40.9), low-prevalence or adult-oriented impulse-control conditions (e.g., F91.0, F63.0), context-specific behavioral diagnoses, (e.g., F42.2, F42.3, F42.4) and remission or mild-state bipolar disorder codes (e.g., F31.11). These exclusions were intended to improve specificity for acute pediatric ED mental health encounters, recognizing that this approach may reduce sensitivity for broader behavioral health burden. Included billing codes are summarized by PECCS category in Table S1.

This study was exempted from ongoing review by the Cincinnati Children’s Institutional Review Board (protocol 2025-0001), and a waiver from the requirement to obtain authorization for use of protected health information was issued for secondary research activities. No patients or families were contacted, and no new patient data were collected.

We considered the earliest signed, “Progress” or “Psychiatry Progress” note that contained both phrases “division of psychiatry” and “intake assessment.” Psychiatric intake assessments are routinely completed during psychiatric evaluation in an emergency setting and contain narrative documentation of recent psychosocial stressors, school functioning, sleep, behavioral issues, and safety concerns. We sought to identify measures that were (1) relevant to acute pediatric mental health crises, (2) not reliably available in structured electronic health records fields or smart forms, (3) sufficiently prevalent for analysis (> 5%), and (4) plausibly actionable for research or intervention planning.

Prompt development used three rounds of 20 notes each. CB, CH, and EM participated in the first two rounds of prompt engineering. JB reviewed a third set of 20 notes as an independent rater who had not participated in prompt development. These prompt-engineering notes were not included in the final agreement results.

We ultimately selected five measures that met our criteria and demonstrated acceptable reliability among human raters: (1) peer conflict (interpersonal issues at school, online, or in another setting), (2) sleep (disruptions or changes to schedule), (3) school academic issues, (4) school attendance issues (absenteeism/truancy), and (5) school discipline issues. Our prompt (Supplement) asked each rater to use “TRUE” or “FALSE” to answer if the patient had recently experienced each measure, with specific instructions to return “NULL” if the note did not provide enough information to determine presence or absence. In addition to the three possible levels, we also considered combining “FALSE” and “NULL” into one category to evaluate binary agreement.[11]

We used an institutional instance of Open WebUI with Ollama to query open-weight large language models through a chat application programming interface. We considered models that could be hosted on premises and excluded models that did not reliably return valid structured JSON without additional completion attempts.[14] The final extraction model family was “gpt-oss”, a set of open-weight mixture-of-experts transformer models released under an Apache 2.0 license. We tested both the 20-billion-parameter and 120-billion-parameter variants and used gpt-oss:120b for all reported results. The model family was selected because it supported local deployment and produced structured outputs for long psychiatric intake notes. The final extraction prompt (see Supplement for full prompt in markdown format) was zero-shot, with no labeled examples in the prompt, and each production note was submitted in a single model call. Each note was used with the prompt for one chat completion with a temperature of zero (a generation setting that minimizes random token sampling by favoring the highest-probability next tokens) and the default context window (128,000 tokens; roughly equivalent to 96,000 words). Because model outputs can still vary across repeated calls even at temperature 0, we separately evaluated stability in the validation set as the percentage prevalence of repeated-queries that were the majority response.

For evaluation, four human raters and the LLM reviewed an additional set of 50 notes from 50 randomly chosen encounters, with only one encounter per patient. Human raters included a mixed group of informaticians, clinicians, and trainees, received the same written extraction definitions as the LLM, and rated notes independently without access to model outputs. We calculated Fleiss’ kappa (κ) to compare inter-rater reliability among human-only ratings and among human plus LLM ratings. Confidence intervals for the paired difference in κ were estimated using 1,000 bootstrap resamples. Using the same 50-note validation set, we calculated the stability of model responses per measure as the percentage of the majority response among 50 independently repeated responses. We also linked note-measure stability values with LLM-human agreement across the 250 note-measure combinations (50 notes × 5 measures), summarized agreement (as percent with human raters) by stability categories (deciles between 80 and 100%), and estimated Pearson correlation and a descriptive linear slope between stability and agreement. After validation, we applied the final prompt to all eligible psychiatric intake assessment notes and summarized the prevalence and intersections of extracted psychosocial variables.

## RESULTS

Among 37,315 emergency department admissions related to PECCS mental health billing codes, 22,284 (59.7%) had an eligible Division of Psychiatry intake assessment note linked to the admission. Of those with an eligible note, 2,767 (12.4%) had multiple notes. After retaining only the first note for each admission, we excluded an additional 3,385 notes. When comparing admissions with and without a linked intake assessment note, we found no differences in gender of patient and small differences in age of patient and date of encounter (Table S2). Further, some diagnostic categories were more common among admissions with notes versus without notes (suicidality and intentional self-inflicted injury: 53% versus 36%, depression: 43% vs 30%) while others were less common (anxiety: 11% vs 22%; Table S2). Intake assessment notes had an average of 1,868 words (median: 1,867; interquartile range: [1,639, 2,105]).

In the 50-note evaluation set, agreement patterns were similar when Fleiss’ κ was calculated among human raters only and when the LLM was included as an additional rater (Figure 1; Tables S3, S4). Agreement using the 20-billion-parameter model was similar compared to the 120-billion-parameter model (Tables S3, S4). Wald tests indicated no statistically significant differences between human-only and human plus LLM κ estimates for any of the five measures in either the three-level coding scheme or the two-level coding scheme. Agreement was highest for sleep disruption (κ, 0.92–0.94) and lowest for academic school issues (0.56–0.71), but generally higher in the two-level coding scheme that combined explicit absence and no evidence. School attendance, school disciplinary issues, and school academic issues showed lower three-level agreement.

**Figure 1:**
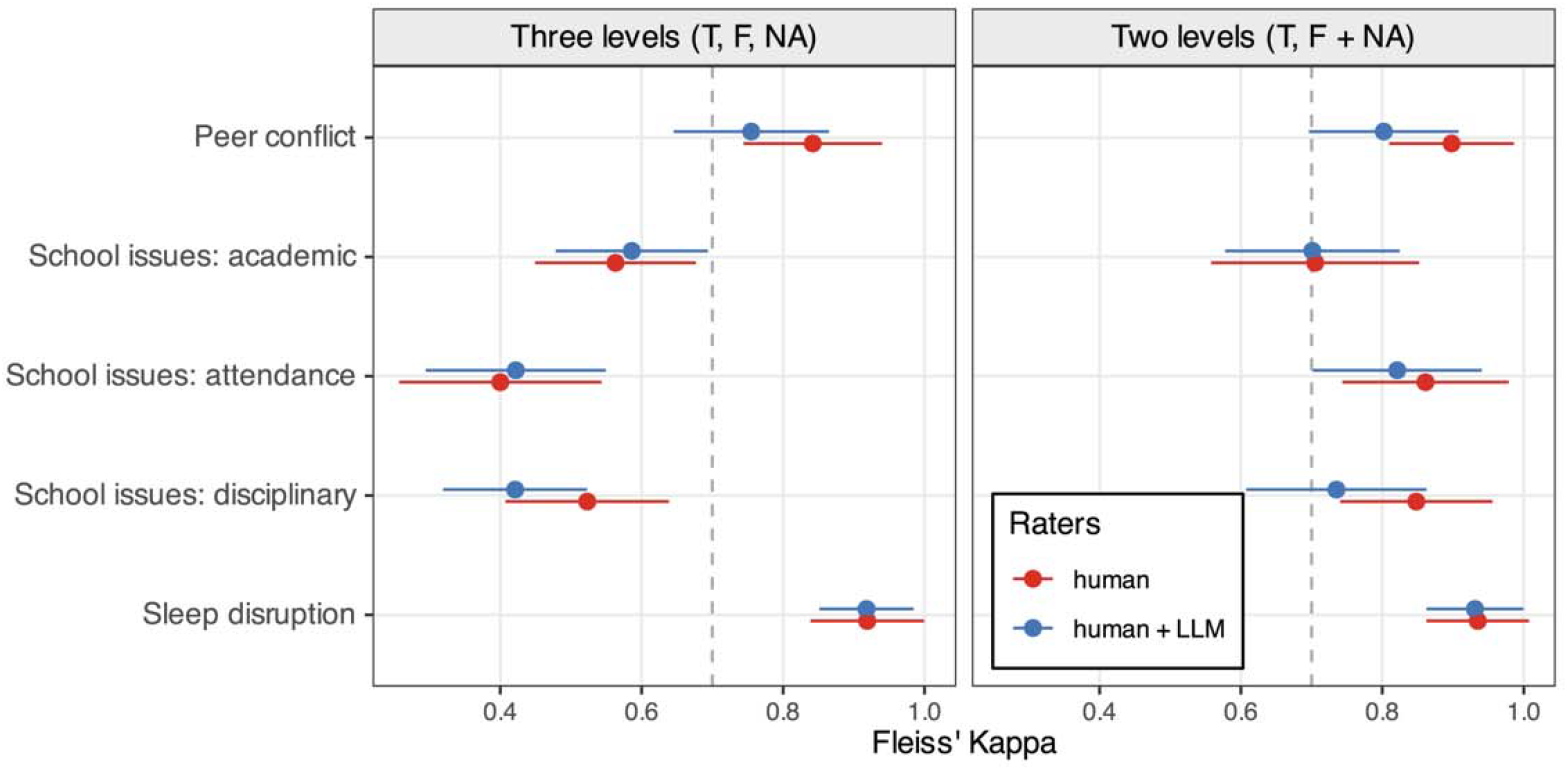
Interrater reliability for humans or humans and LLM across five measures extracted from psychiatric intake assessment notes using a three-level (true, false, null) and two-level (true, false or null) coding schemes. Alt text: Forest plots compare Fleiss’ kappa estimates and confidence intervals for human-only raters and human plus LLM raters across five psychosocial measures under three-level and two-level coding schemes.

Across the 50-note evaluation set, the LLM produced stable results when queried repeatedly (Table S5), with mean stability ranging from 92.5% in peer conflict to 98.6% in sleep issues. Sleep and academic school issues were the most stable, with 92.0% and 72.0%, respectively, of notes showing 100% stability, and only 2.0% and 0.0%, respectively, of notes having stability <80%. More variation was observed in peer conflict (100% stable: 56.0%; <80% stable: 18.0%), school disciplinary issues (58.0%; 6.0%), and school attendance issues (72.0%;8.0%).

At the note-measure level, lower stability scores were associated with lower agreement with human raters. Across 250 note-measure combinations (50 notes × 5 measures), the average percentage of human ratings the LLM agreed with increased from 42.6% when stability was <80% to 82.7% when stability was 100% (Table S6). Stability was positively associated with LLM-human agreement (Pearson r=0.36), with a descriptive linear slope suggesting that each 10 percentage-point increase in stability was associated with approximately 13 percentage points of additional LLM-human agreement. Agreement patterns varied by measure, but agreement was higher when stability was ≥80% than when stability was <80% for measures with low-stability observations (Table S7).

When processing 22,284 psychiatric intake assessment notes linked with an admission, the LLM returned responses that could not be parsed as JSON for 14 notes (<0.01%), which were excluded from further analysis. Across the 22,270 intake assessment notes with successfully extracted structured data, sleep disruption was the most detected measure (57.7%), and peer conflict (47.2%), school disciplinary (43.3%), and academic (43.4%) measures were also commonly detected (Table 1). The least commonly detected measure was school attendance (16.9%), with most of the responses being indeterminate (81.9%). Sleep disruption and school disciplinary measures were able to be negated more often than the other measures (37.7% and 43.1%, respectively).

**Table 1.** Structured data extracted for 22,270 psychiatric intake assessment notes with successfully parsed LLM output on five measures that could be rated as “TRUE” (detected), “NULL” (indeterminate), or “FALSE” (negated).

| measure | N true | N null | N false |
| --- | --- | --- | --- |
| Peer conflict | 10,503 (47.2%) | 10,825 (48.6%) | 942 (4.2%) |
| Sleep disruption | 12,876 (57.7%) | 1,028 (4.6%) | 8,396 (37.7%) |
| Academic issues | 9,668 (43.4%) | 8,212 (36.9%) | 4,390 (19.7%) |
| Attendance issues | 3,753 (16.9%) | 18,245 (81.9%) | 272 (1.2%) |
| Disciplinary issues | 9,651 (43.3%) | 3,021 (13.6%) | 9,598 (43.1%) |

Figure 2 demonstrates substantial overlap among extracted risk measures. Grouped by intersection, the most common was sleep disruption alone, followed by the absence of all five risk measures (9.4%). Among co-occurring risk profiles, sleep disruptions appeared frequently in combination with peer conflict and school-related academic or disciplinary issues. School attendance issues contributed relatively little to the largest intersections and appeared more often as part of multi-measure combinations than as a large standalone category. The distribution of intersections was highly skewed (Figure 2), with a small number of common profiles and long tail of less frequent combinations, indicating heterogeneity in how risk-related concerns were documented across intake notes.

**Figure 2:**
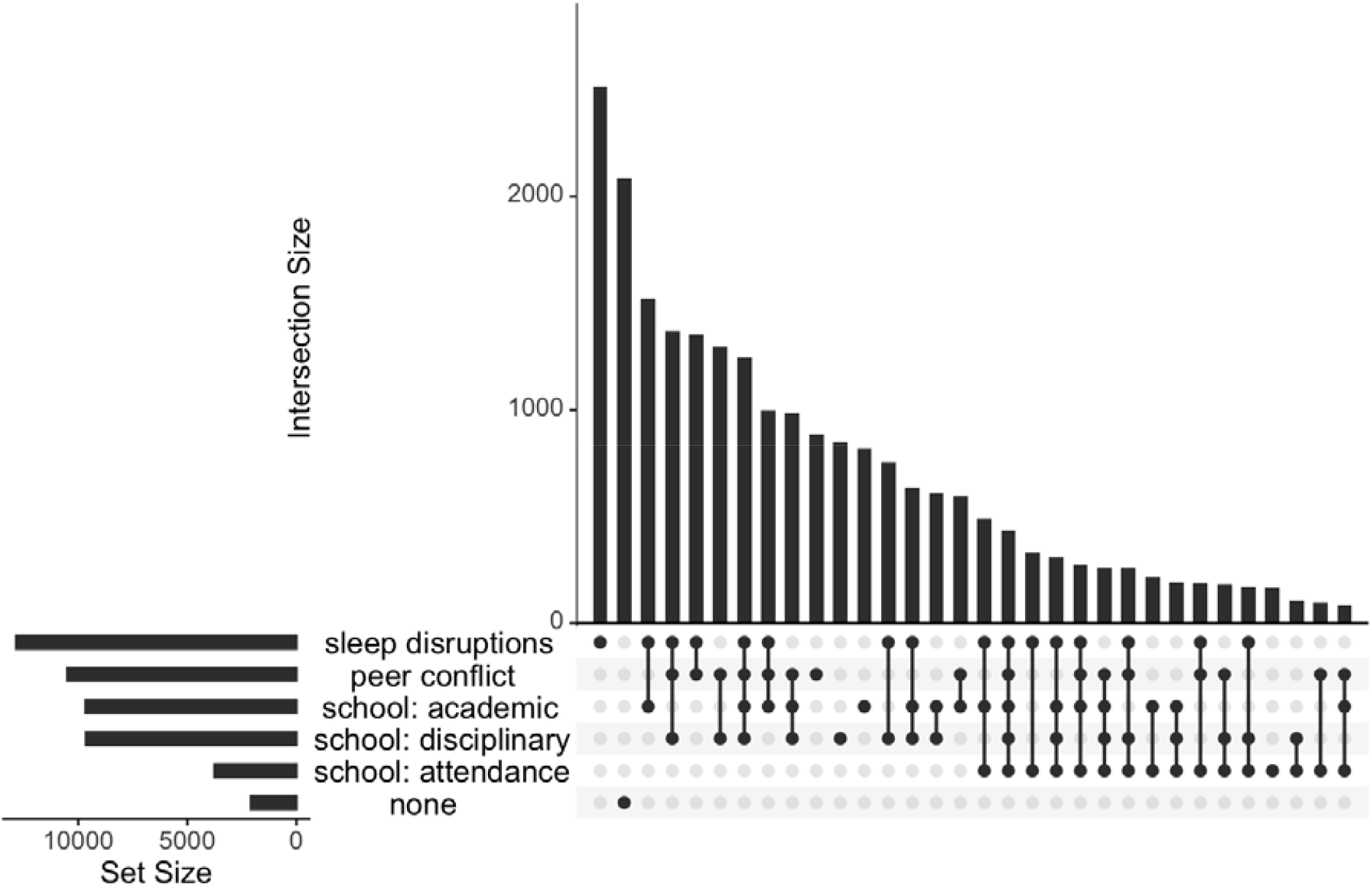
Prevalence of individual and intersecting groups of psychosocial measures detected across psychiatric hospitalizations with an intake assessment. Alt text: UpSet-style plot showing set sizes and intersection counts for documented sleep disruptions, peer conflict, academic issues, disciplinary issues, attendance issues, and no detected measures across psychiatric hospitalizations.

## DISCUSSION

In this single-center cohort of pediatric psychiatric emergency admissions, an on-premises open-weight LLM extracted five psychosocial measures from intake notes with reliability similar to human raters. We developed a methodological process for local LLM extraction from long psychiatric intake notes into structured JSON and used it to find which psychosocial stressors were recently common across psychiatric emergency admissions. For this feasibility study, we considered κ ≥ 0.70 pragmatically acceptable for population-level descriptive extraction after local validation and the two-level coding scheme (true, not true) showed acceptable Fleiss’ κ for all five measures when the LLM was included with human raters. Under the more granular three-level coding scheme (true, false, null), reliability remained acceptable for peer conflict (κ = 0.76) and sleep (κ = 0.92), but fell below the 0.7 threshold for school academic issues (κ = 0.59), school attendance issues (κ = 0.42), and school disciplinary issues (κ = 0.42), suggesting that these school-related measures were less reliable when distinguishing detected, negated, and indeterminate response. After validation, the workflow processed more than 22,000 notes with very few outputs that failed to parse, supporting the feasibility of applying locally hosted LLMs to large EHR-derived psychiatric text.

The difference between the three-level and two-level coding scheme highlights the importance of differentiating explicit absence with lack of documentation. Because of this, extracted prevalence estimates should be interpreted as documented psychosocial concerns rather than definitive measures of whether a stressor was present. Nonetheless, high frequency of sleep disruption, peer conflict, academic issues, and disciplinary issues demonstrates that psychiatric emergency admissions commonly occur in the context of recent functional and interpersonal stressors. Attendance problems were detected less often than the other issues, but the high proportion of indeterminate responses suggests that it may not be consistently assessed or documented. The measure-specific reliability patterns suggest that some psychosocial constructs are more readily recoverable from psychiatric intake documentation than others. Sleep disruption and peer conflict may be documented in more direct language, whereas school-related measures often require interpretation across academic performance, attendance, behavior, school avoidance, and discipline context.

Human agreement was also lower for some school-related measures, suggesting that the extraction task uncovered ambiguity in the underlying documentation and construct definitions. Future work could benefit from more explicit annotation prompts or pre-labeled examples, but our results suggest that a simple, zero-shot approach can be useful. Indeed, the use of an on-premises open-weight model reduced privacy and governance barriers and allowed us to define measures, prompts, and output schemas that best match our own documentation practices and research questions.

Our study builds on emerging work by demonstrating that LLMs can support pediatric, psychiatric clinical information extraction by reasoning about locally defined psychosocial constructs from long (often thousands of words) intake assessments instead of classifying short snippets. However, extracted measures remain vulnerable to documentation bias (e.g., clinician personal characteristics, caregiver availability, acuity, language, race and ethnicity, insurance status); these biases and changes in intake templates and standard operating procedures over time may all influence whether a psychosocial stressor appears in a note. More broadly, psychiatric intake notes are clinical documents created for care rather than research. Extracted measures may reflect patient or caregiver report, clinician interpretation, copied or templated text, and the acute crisis context. Thus, measures should be treated as derived documentation-based variables rather than independently verified exposures.

A notable feature of our approach is that we leveraged LLM non-determinism rather than attempting to eliminate it. Prior work has shown that LLM outputs may vary across repeated calls even under settings intended to be deterministic, and that agreement across repeated calls can be informative for prediction confidence or factual reliability.[15] The stability analysis found that when repeated model queries converged, outputs were more likely to agree with human ratings, suggesting that repeated-query stability may help flag lower-confidence note-measure classifications for review in future workflows.

Our validation sample provides initial evidence of feasibility but was not large enough to fully evaluate performance across demographic, diagnostic, temporal, or documentation subgroups. In addition, because agreement was measured against human raters rather than an adjudicated clinical gold standard, these results show that the model interpreted documentation similarly to reviewers, not that it established the true presence or absence of each stressor. Further, because only 59.7% of PECCS mental health admissions had a linked intake note, and diagnostic categories differed between admissions with and without notes, the findings are most generalizable to encounters receiving documented psychiatric intake assessment rather than to all pediatric mental health-related emergency admissions. One operational explanation is that encounters requiring medical stabilization before psychiatric assessment may not consistently follow the same intake documentation workflow, which could contribute to differential note availability across diagnostic categories. Additionally, our cohort definition may include presentations for medical complaints that did not receive a psychiatric assessment but did result in a psychiatric diagnosis. Indeed, the goal of the psychiatric intake assessment is safety, and it is focused on suicidality, homicidality, and self-harm. Although generalizability of our findings is limited, the overall approach of local validation followed by structured extraction from narrative notes is portable to other institutions if prompts, models, and target measures are re-evaluated in each setting.

## CONCLUSION

Locally hosted open-weight LLMs may provide a feasible and accessible method for transforming pediatric psychiatric intake documentation into structured research variables at scale. These tools can help characterize the psychosocial context of psychiatric emergency admissions, but their outputs should be interpreted as documented stressors and validated carefully before use in clinical decision-making or population-level inference.

## Data Availability

The data underlying this article cannot be shared because they contain protected health information under HIPAA and were used under an IRB-approved research study with exempt use of protected health information and no sharing beyond the study investigators.

## Acknowledgements

We would like to acknowledge the support of the Informatics Shared Facility in Information Services for Research (IS4R) at Cincinnati Children’s Hospital Medical Center (RRID:SCR_022622).

