## Supplemental Materials for "Characterizing Documented Psychosocial Stressors in Pediatric Psychiatric Emergencies with an Open-Weight Large Language Model"

**Supplement**

Prompt

```
You are a JSON extraction assistant.
Output only a single JSON object (no prose, no markdown, no explanations).
Given a clinical psychiatric intake note, extract the following fields:

- `peer_conflict`
  - description: Has the patient recently had peer conflict issues at school, online, or in another setting (verbal, physical, or social)?
  - type: boolean, null
- `sleep`
  - description: Has the patient recently experienced sleep disruptions or changes to their sleep schedule?
  - type: boolean, null
- `school_issues_academic`
  - description: Has the patient experienced academic struggles at school?
  - type: boolean, null
- `school_issues_attendance`
  - description: Has the patient had issues with absenteeism or truancy from school?
  - type: boolean, null
- `school_issues_disciplinary`
  - description: Has the patient experienced disciplinary problems at school?
  - type: boolean, null

For boolean data, return `false` if the note explicitly indicates absence of the risk factor and return `null` if the note does not provide enough information to determine presence or absence.
In addition to each field value, concisely provide evidence used to determine the value. Make each field have fields `value` and `evidence`.
```

Table S1

Select billing codes used to define pediatric psychiatric exacerbations grouped by the corresponding PECCS category. Examples of excluded codes for impulse control disorders: "pathological gambling", "kleptomania", "trichotillomania"; for anxiety disorders: "hoarding disorder", "excoriation disorder", "nonpsychotic mental disorder")

| **PECCS category** | **ICD-10-CM codes** |
| --- | --- |
| Adjustment disorders | F43.20, F43.21, F43.22, F43.23, F43.24, F43.25, F43.29, F43.8, F43.9 |
| Anxiety disorders | F41.0, F41.1, F41.3, F41.8, F41.9, F43.0 |
| Disorders usually diagnosed in infancy childhood or adolescence | F93.0 |
| Attention-deficit hyperactivity disorder | F90.0, F90.1, F90.2, F90.8, F90.9 |
| Conduct disorder | F91.1, F91.2, F91.8, F91.9 |
| Oppositional defiant disorder | F91.3 |
| Disruptive behavior disorder | R46.89 |
| Impulse control disorders NEC | F63.1, F63.89, F63.9, R45.850 |
| Intermittent explosive disorder | F63.81 |
| Mood disorders | F34.0, F34.1, F34.8, F34.81, F34.89, F34.9, F39 |
| Mood disorders (bipolar disorder) | F31.0, F31.10, F31.12, F31.13, F31.2, F31.30, F31.31, F31.32, F31.4, F31.5, F31.60, F31.61, F31.62, F31.63, F31.64, F31.70, F31.73, F31.75, F31.77, F31.81, F31.89, F31.9 |
| Mood disorders (major depressive disorder) | F32.0, F32.1, F32.2, F32.3, F32.5, F32.8, F32.89, F32.9, F33.0, F33.1, F33.2, F33.3, F33.40, F33.8, F33.9 |
| Mood disorders (manic episodes) | F30.2, F30.8, F30.9 |
| Personality disorders | F60.3, F60.89, F60.9 |
| Phobias | F40.00, F40.01, F40.10, F40.11, F40.298 |
| Posttraumatic stress disorder | F43.10, F43.11, F43.12 |
| Schizophrenia and other psychotic disorders | F20.0, F20.1, F20.2, F20.3, F20.5, F20.81, F20.89, F20.9, F22, F23, F24, F25.0, F25.1, F25.8, F25.9, F29 |
| Suicide and intentional self-inflicted injury | R45.851, T14.91, T36.1X2A, T37.5X2A, T38.1X2A, T38.3X2A, T39.012A, T39.1X2A, T39.1X2D, T39.312A, T39.392A, T39.8X2A, T39.92XA, T40.2X2A, T40.4X2A, T40.7X2A, T40.8X2A, T42.4X2A, T42.6X2A, T42.8X2A, T43.012A, T43.212A, T43.222A, T43.292A, T43.3X2A, T43.592A, T43.612A, T43.622A, T43.632A, T44.3X2A, T44.7X2A, T44.992A, T45.0X2A, T45.2X2A, T45.4X2A, T46.1X2A, T46.4X2A, T46.5X2A, T47.0X2A, T47.92XA, T48.1X2A, T48.3X2A, T48.4X2A, T49.0X2A, T50.902A, T50.992A, T51.0X2A, T54.1X2A, T54.92XA, T65.892A, T71.162A |

Table S2

Differences in patient- and encounter-level characteristics by whether or not an associated psychiatric intake assessment note was present in the electronic health record. Significant differences were assessed using p-values from Chi-squared statistic for categorical characteristics (specifically applying a Bonferroni correction for tests on billing code categories) and a Wilcoxon rank-sum statistics for continuous characteristics.

| Characteristic | No note (n=15031) | Note (n=22,284) | stat sig (p<0.05) |
| --- | --- | --- | --- |
| Age (years) | 14.4 | 14.5 | * |
| Gender (% female) | 57.2 | 58.1 |  |
| Encounter date | 2020-10-07 | 2020-07-11 | * |
| Adjustment disorders (%) | 8.3 | 9.5 | * |
| Anxiety disorders (%) | 21.7 | 11.5 | * |
| Disorders usually diagnosed in infancy childhood or adolescence (%) | 0.2 | 0.1 |  |
| Impulse control disorders nec (%) | 4.4 | 7.8 | * |
| Intermittent explosive disorder (%) | 10.4 | 17.5 | * |
| Mood disorders (%) | 13.8 | 22.6 | * |
| Mood disorders bipolar disorder (%) | 4.1 | 5.0 | * |
| Mood disorders major depressive disorder (%) | 30.1 | 43.2 | * |
| Mood disorders manic episodes (%) | 0.1 | 0.1 |  |
| Personality disorders (%) | 0.9 | 1.2 |  |
| Phobias (%) | 0.6 | 1.0 | * |
| Posttraumatic stress disorder (%) | 11.9 | 16.2 | * |
| Schizophrenia and other psychotic disorders (%) | 2.8 | 3.7 | * |
| Suicide and intentional self inflicted injury (%) | 36.4 | 53.2 | * |

Table S3

Fleiss’ kappa (κ), a measure of interrater reliability for human-only raters or human and LLM raters (120b and 20b models) across five measures extracted from psychiatric intake assessment notes using a two-level coding scheme (detected vs. negated or indeterminate).

| Measure | Human κ (95% CI) | Human + LLM-120b κ (95% CI) | Human + LLM-20b κ (95% CI) |
| --- | --- | --- | --- |
| Peer conflict | 0.90 (0.81, 0.99) | 0.80 (0.70, 0.91) | 0.81 (0.70, 0.91) |
| School issues: academic | 0.71 (0.56, 0.85) | 0.70 (0.58, 0.82) | 0.74 (0.61, 0.86) |
| School issues: attendance | 0.86 (0.74, 0.98) | 0.82 (0.70, 0.94) | 0.78 (0.66, 0.91) |
| School issues: disciplinary | 0.85 (0.74, 0.96) | 0.74 (0.61, 0.86) | 0.71 (0.58, 0.84) |
| Sleep disruption | 0.94 (0.86, 1.0) | 0.93 (0.86, 1.0) | 0.91 (0.84, 0.99) |

Table S4

Fleiss’ kappa (κ), a measure of interrater reliability for human-only raters or humans and LLM raters (120b and 20b models) across five measures extracted from psychiatric intake assessment notes using a three-level coding scheme (detected vs. negated vs. indeterminate).

| Measure | Human κ (95% CI) | Human + LLM-120b κ (95% CI) | Human + LLM-20b κ (95% CI) |
| --- | --- | --- | --- |
| Peer conflict | 0.84 (0.74, 0.94) | 0.76 (0.65, 0.87) | 0.76 (0.65, 0.87) |
| School issues: academic | 0.56 (0.45, 0.68) | 0.59 (0.48, 0.69) | 0.61 (0.51, 0.72) |
| School issues: attendance | 0.40 (0.26, 0.54) | 0.42 (0.30, 0.52) | 0.40 (0.28, 0.53) |
| School issues: disciplinary | 0.52 (0.41, 0.64) | 0.42 (0.32, 0.52) | 0.43 (0.33, 0.53) |
| Sleep disruption | 0.92 (0.84, 1.0) | 0.92 (0.85, 0.99) | 0.87 (0.79, 0.96) |

Table S5

Stability (% of 50 repeated responses that agreed with the most common response) was characterized for each measure across all 50 notes in the evaluation set.

| Measure | Mean stability | % stability <80% | % stability =100% |
| --- | --- | --- | --- |
| Peer conflict | 92.5% | 18% | 56% |
| School issues: academic | 97.7% | 0% | 72% |
| School issues: attendance | 96.2% | 8% | 72% |
| School issues: disciplinary | 94.1% | 6% | 58% |
| Sleep disruption | 98.6% | 2% | 92% |

Table S6

LLM-human agreement by stability band across 250 note-measure combinations in the 50-note evaluation set.

| Stability band | n note-measures | Mean stability | Mean LLM-human agreement | LLM agrees with human plurality |
| --- | --- | --- | --- | --- |
| <80% | 17 | 65.2% | 42.6% | 47.1% |
| 80–89% | 18 | 84.4% | 58.3% | 60.0% |
| 90–99% | 40 | 95.6% | 69.4% | 71.8% |
| 100% | 175 | 100.0% | 82.7% | 88.9% |

Table S7

Measure-specific stability and LLM-human agreement across the 50-note evaluation set.

| Measure | Mean stability | n with stability <80% | % with 100% stability | Overall LLM-human agreement | LLM-human agreement when stability <80% | LLM-human agreement when stability ≥80% |
| --- | --- | --- | --- | --- | --- | --- |
| Peer conflict | 92.5% | 9 | 56.0% | 86.5% | 61.1% | 92.1% |
| School issues: academic | 97.7% | 0 | 72.0% | 78.5% | — | 78.5% |
| School issues: attendance | 96.2% | 4 | 72.0% | 69.0% | 43.8% | 71.2% |
| School issues: disciplinary | 94.1% | 3 | 58.0% | 52.5% | 0.0% | 55.9% |
| Sleep disruption | 98.6% | 1 | 92.0% | 94.0% | 0.0% | 95.9% |
